# Multi-stage reweighting to correct for participation bias in a nationwide biobank with nested recruitment

**DOI:** 10.64898/2026.04.01.26349852

**Authors:** Jacob Træholt, Maria Didriksen, Dorte Helenius, Lea A. N. Christoffersen, Khoa M. Dinh, Joseph Dowsett, Christina Mikkelsen, Lotte Hindhede, Liam J. E. Quinn, Mie T. Bruun, Bitten Aagaard, Thomas F. Hansen, Henrik Hjalgrim, Klaus Rostgaard, Erik Sørensen, Christian Erikstrup, Ole B. V. Pedersen, Torben Hansen, Andrew J. Schork, Bo Markussen, Sisse R. Ostrowski

**Author notes:** **Address for correspondence:** Jacob Træholt, MSc, PhD Student, Department of Clinical Immunology, Rigshospitalet, Copenhagen University Hospital, Copenhagen, Denmark.

## Abstract

Selective participation in biobanks often compromises inference to the general population, particularly when selection occurs across multiple stages, whether at recruitment or during subsequent participation. Inverse probability (IP) weighting can reduce systematic differences using suitable external benchmarks, but most applications assume a single selection process. Here, we present a multi-stage IP-weighting framework and apply it to the Danish Blood Donor Study (DBDS), a nationwide biobank embedded in Denmark’s blood-donation infrastructure. Using national registers, we estimated year-specific probabilities of (i) donation activity and (ii) DBDS enrolment conditional on donation activity, yielding two-stage inclusion weights for 169,893 participants. These weights reduced inclusion-associated imbalance across the 52 auxiliary variables in the probability models by 97.6% (median) and, despite strong health selection under donation-based recruitment, reduced relative-prevalence discrepancies across held-out prescription phenotypes by 69.7% (median). The effective sample size after weighting was 30,627 (18.0% of 169,893). Combining the inclusion weights with questionnaire-specific response weights across five DBDS questionnaires (>500 questions) produced the largest changes from unweighted to weighted responses for health behaviours and symptom severity, including tobacco and alcohol consumption, menstrual-pain severity, restless-legs severity, nocturia, sleep disturbance, and fatigue. These findings support multi-stage IP-weighting to improve population alignment in biobanks with staged selection.

## Introduction

Large volunteer-based cohort studies and biobanks are important resources for biomedical research when linked to self-report data, population registers, and electronic health records^1^, yet the processes by which individuals enter and continue to participate may compromise inference. When participation depends on exposures, outcomes, or their determinants, the resulting study population will typically differ systematically from the population to which inference is intended. This participation bias often manifests as a healthy volunteer pattern^2,3^, with participation tending to correlate with sociodemographic position, diagnostic history, and health-related behaviours. Moreover, conditioning on study participation can introduce collider bias by opening noncausal paths when participation is influenced by both exposure and outcome^4,5^, thereby distorting associations in both magnitude and direction. As a result, naive analyses may fail to generalize to the target population, limiting external validity and population-level interpretability^2,3^.

Recent studies in UK Biobank (UKB)^6,7^ have applied inverse probability (IP) weighting to correct for participation bias by estimating each participant’s probability of inclusion using predictors measured in both UKB and external reference data. Participants are then weighted by the inverse probability to construct a pseudo-population that closely reflects the intended target population, up-weighting under-represented and down-weighting over-represented individuals. Using representative reference surveys or census microdata, IP-weighting markedly reduced inclusion-related differences and recovered population-level distributions for key characteristics^8,9^. Reweighting also altered downstream genetic analyses, shifting single-nucleotide polymorphism effect sizes and, in some instances, the set of associations detected, despite a reduced effective sample size^8,10^. Together, these studies establish IP-weighting as a feasible and effective strategy for correcting healthy-volunteer participation bias in UKB and provide a framework that can be adapted to other volunteer-based biobanks, contingent on the availability of appropriate external benchmarks. However, these applications focused primarily on modelling initial study inclusion, leaving subsequent participation stages, such as questionnaire response, less well explored.

The Danish Blood Donor Study (DBDS)^11,12^, which includes extensive genetic, biomarker, and questionnaire data, provides a distinct setting in which to extend this approach. In contrast to UKB, which recruited middle-aged adults through a time-fixed invitation campaign (2006–2010), DBDS has enrolled more than 170,000 adult blood donors continuously across Denmark’s nationwide blood-donation infrastructure since 2010. Recruitment is intrinsically two-stage: individuals must first present and be deemed eligible to donate, and only then can they be invited to participate in DBDS. This nested recruitment structure yields a particularly healthy subset of the Danish adult population^13^ and reflects donor eligibility screening followed by study consent. In Denmark, comprehensive national registers offer near-complete sociodemographic, diagnostic, and prescription coverage^14^ that can be used as auxiliary variables for modelling participation. Although registers lack some lifestyle and self-reported measures obtainable from surveys, their accuracy and population coverage make them well suited to this purpose.

In this paper, we develop and apply an IP-weighting framework aligned with DBDS’s nested two-stage recruitment process. For each inclusion year, we estimate (i) the probability that an adult in the Danish resident population has at least one donation event and (ii) the probability of enrolling in DBDS conditional on such an event. Each probability is estimated using LASSO-regularized logistic regression with register-based auxiliary variables, and their product yields an individual inclusion probability from which we construct IP-weights. We then evaluate the weighted DBDS cohort against the corresponding target population using a broad set of sociodemographic and health-related variables, including both variables used in the propensity models and held-out prescription phenotypes. Finally, we demonstrate how the framework can be extended with a third, questionnaire-response stage to model participation in five DBDS questionnaires focusing on lifestyle and self-reported health. Although motivated by DBDS, the framework applies to other biobanks and cohort studies with staged selection when suitable external benchmarks are available.

## Methods

We used national registers to construct two-stage inclusion weights for selection into DBDS and extended this framework with a third stage for questionnaire response. Our target population was the Danish resident population eligible for DBDS enrolment from 1 March 2010 to 31 December 2024, with eligibility defined here as being within the year-specific age range for at least one day during the calendar year (Supplementary Table S1) and restricted to individuals without prior DBDS enrolment.

### The Danish Blood Donor Study

DBDS is an ongoing nationwide cohort and biobank embedded within the Danish blood bank infrastructure^11,12^. Since 1 March 2010, adult blood donors have been invited during routine donation visits to provide written informed consent and complete a baseline questionnaire, including permission for research use of archived plasma samples collected at every donation, both historically and prospectively. Approximately every two years, DBDS introduces a new questionnaire administered at donation visits, covering lifestyle and other health-related questions (DBDS1–DBDS5 in the present work). Moreover, online questionnaires on targeted health topics are periodically distributed via secure digital mail to all consenting participants, regardless of current donor status. In addition, DBDS includes biomarker panels in nested sub-cohorts, genome-wide genotyping for a substantial subset of participants^15,16^, and is linked to Danish demographic, socioeconomic, hospital diagnosis, and prescription registers via the unique personal identifier assigned to all Danish residents^17^.

### Auxiliary variables

We selected auxiliary variables from national registers^14^ to capture key domains plausibly related to blood donation and selection into DBDS, guided by prior evidence and register availability. For each DBDS inclusion year, we derived 52 variables that were consistently defined for all individuals in the corresponding risk set (described below) and measured as of 31 December of the preceding year. These variables encompassed: (legal) sex, age, civil status, region of residence, country of origin, immigrant status, household size, household composition, number of children, highest attained education, employment status, profession, income percentile (computed each year across all 17–74 year-olds), number of unique redeemed prescriptions in the previous year, number of unique hospital diagnoses in the previous three years, Charlson comorbidity index, indicator functions for all first-level ATC (Anatomical Therapeutic Chemical) groups and ICD-10 (International Classification of Diseases, 10th Revision) chapters, as well as sampling year (calendar year of the risk set).

To allow flexible modelling of selection probabilities, we included all two-way interactions after categorizing and coding variables as described in Supplementary Table S2, with categorical variables dummy-coded using a reference category. This resulted in a preliminary set of 6,443 predictors (excluding within-variable interactions). For brevity, we refer to the resulting design matrix as *X*. Several predictors capture overlapping constructs; correlations were therefore expected and accommodated through regularization in the two-stage selection model described below. No donation-history or visit-level variables were included to retain interpretability and overlap with the target population.

### Two-stage selection model and estimation

For each calendar year in the study period, from 1 March 2010 to 31 December 2024, we constructed a risk set comprising all individuals who were alive and registered as residents in Denmark on 31 December of the preceding year, met the age-range criterion for DBDS enrolment, were not already enrolled in DBDS, and could in principle (not accounting for deferrals) have had a donation event. We considered only donation visits of types that were used for DBDS enrolment during the study period, primarily whole-blood, plasma, and platelet donations.

Within each annual risk set, we then defined two nested indicators of donation activity and DBDS enrolment:

- **Stage 1 (donation activity)**: 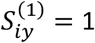 if individual *i* had at least one registered donation in year 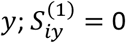 otherwise.
- **Stage 2 (DBDS enrolment)**: 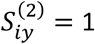 if individual *i* enrolled in DBDS at any donation visit in year 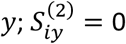 otherwise.

For an individual *i* in the annual risk set for year *y*, let *X*_*iy*_ denote the vector of auxiliary variables measured on 31 December of the year preceding *y*. We model the probability of DBDS inclusion in year *y* as a product of two conditional probabilities,

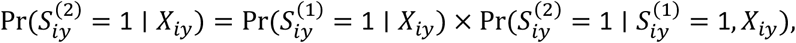

where the first factor models selection from the annual risk set into the active donor pool, and the second factor models selection from that active donor pool into DBDS.

For each DBDS participant enrolled in year *y*, we next obtained the model-based inclusion probability 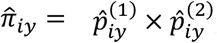, where 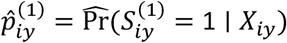 and 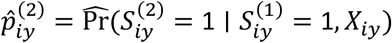 were estimated from the stage 1 and stage 2 propensity models, respectively. We estimated the stage-specific propensities using logistic regression with LASSO regularization and five-fold cross-validation. For each stage, we pooled all annual risk sets (2010–2024) and assigned individuals to five out-of-fold groups, stratified by inclusion year so that each year contributed approximately equal proportions to each fold.

For stage 1, the outcome was 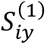 for all risk-set individuals. To reduce computational burden, we included all donors and a 5% simple random sample of non-donors for each year. In the model fit, donors received weight 1 and sampled non-donors weight 1/0.05 to account for subsampling. For stage 2, the outcome was 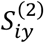 among individuals with at least one registered donation in year *y*.

### Weight construction and calibration

For each stage *s* and inclusion year *y*, we defined stabilized weights 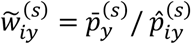, where 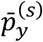 is the observed marginal fraction of 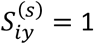 in year *y*. The stage-specific stabilized weights were then multiplied to form 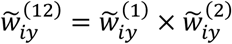, and the resulting weights were winsorized across all years at the 1st and 99th percentiles to limit the influence of extreme weights^18–21^. Year-specific stabilization makes weight scales more comparable across years and reduces the extent to which pooled winsorization is driven by calendar years with low or high selection fractions.

To further improve marginal alignment with the target population, we calibrated 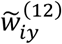 to external margins using iterative proportional fitting (raking)^22–24^, which adjusts weights iteratively so weighted DBDS totals match population totals for the selected margins. Calibration targets were chosen from the auxiliary variables through a forward-selection procedure: starting from a core set of margins (sex × age and sampling year), we added margins one at a time to improve auxiliary-variable balance while limiting reductions in effective sample size; the selected margins are listed in Supplementary Table S3. The final inclusion weights were then rescaled to have mean 1,

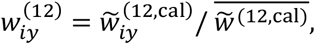

with 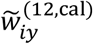 denoting the calibrated weights. The effective sample size was subsequently computed as 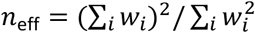, where 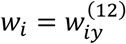.

### Evaluation of weighting performance

We first evaluated the inclusion weights by quantifying how strongly DBDS inclusion remained associated with auxiliary variables after weighting. For each main-effect auxiliary variable, we fitted univariable logistic regression models for DBDS inclusion in the pooled annual population risk set using a quasi-binomial family with a logit link to accommodate potential overdispersion, once unweighted and once after applying the calibrated inclusion weights. DBDS participants in their most recent risk set were assigned their inclusion weight, 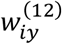, whereas all other individuals (including DBDS participants in earlier risk sets) were assigned weight 1. Standardized regression coefficients and 95% Wald confidence intervals (CIs) were estimated; attenuation toward zero after weighting indicates reduced predictability of inclusion from the observed variables. We summarized attenuation as the relative change in absolute standardized coefficients, 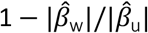, where 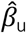 and 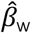 denote the unweighted and weighted coefficients, respectively; absolute values were used so attenuation reflects changes in magnitude irrespective of sign.

For presentation of coefficient estimates, we used a prespecified subset of auxiliary variables spanning age, sex, income percentile, educational attainment, employment status, household composition, immigrant status, number of unique redeemed prescriptions in the previous year, and number of unique hospital diagnoses in the previous three years. For this subset, we additionally compared weighted and unweighted means and standard deviations (SDs) in DBDS with those in the target population. Weighted means and SDs were computed as 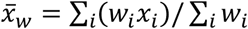 and 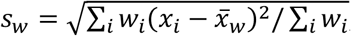, respectively.

To assess generalization beyond the auxiliary-variable set used for propensity estimation and calibration, we evaluated the weighting performance using held-out prescription phenotypes. Specifically, we compared target-population prevalences with unweighted and weighted DBDS prevalences for 3-character ATC groups, defining ATC-group use as ≥1 redeemed prescription in the 1-year window ending on the risk-set date (31 December). We restricted the analysis to ATC groups with ≥500 DBDS participants. These 3-character ATC groups were not included directly in the propensity models, which used only first-level ATC groups, thereby assessing generalization to more granular prescription phenotypes. For each ATC group, we quantified deviation from target-population prevalence using the absolute log prevalence ratio, | log(*p*_DBDS_/*p*_target_) |, which treats over- and underestimation symmetrically on a multiplicative scale, and summarized whether weighting moved the DBDS prevalence closer to the target-population prevalence.

### Extension to questionnaire-response weighting

For DBDS questionnaires 1–5, we introduced a third stage to account for non-response among DBDS participants. For each questionnaire *q*, we defined 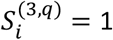 if participant *i* contributed responses to questionnaire *q*, and 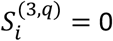 otherwise. The questionnaire-specific risk sets comprised DBDS participants who were alive, resident in Denmark, and eligible for invitation at any time during the questionnaire’s field period.

We constructed an auxiliary-variable vector 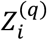 as follows: if participant *i* enrolled in DBDS before questionnaire *q* started, 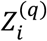 was based on register-based variables measured on 31 December of the year preceding questionnaire launch; if enrolment occurred during the questionnaire period, 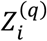 was defined as *X*_*iy*_ from the year preceding inclusion. In both cases, the design matrix for stage 3 included the same main effects and two-way interactions as used in the inclusion models.

Within each questionnaire risk set, we fitted a logistic LASSO regression model with five-fold cross-validation to estimate the probability of 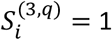, and constructed questionnaire-specific stabilized weights (across all response-years) using the observed response fraction. For analyses of questionnaire *q*, we combined the non-winsorized inclusion weights with the stabilized stage-3 weights, 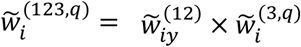. We then winsorized and calibrated 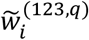 using the same winsorization percentiles and the same types of population margins as for the two-stage inclusion weights, except that sampling year was omitted for calibration, and rescaled the resulting questionnaire-specific weights to mean 1 among respondents. These three-stage weights approximate the inverse of the model-based probability that an individual in the defined risk sets enters DBDS and subsequently contributes responses to questionnaire *q*.

## Results

### Study population and annual risk sets

We identified 172,562 adults enrolled in DBDS as of 31 December 2024. Of these, 169,893 participants with complete register-based auxiliary variables available as of 31 December of the calendar year preceding enrolment comprised the analytic study population. For each inclusion year in the study period (from 1 March 2010 to 31 December 2024), we defined a corresponding annual population risk set of individuals who were alive and resident in Denmark at the end of the preceding year, met the age criteria, and had not already enrolled in DBDS. Annual risk sets comprised approximately 3.5–4.1 million individuals and formed the basis for estimating year-specific inclusion probabilities.

Across the study period, annual DBDS enrolment was highest in the earliest years, declined markedly after the first three years, and subsequently fluctuated with alternating increases and decreases, including a pronounced dip around 2020 (8 months without inclusion due to COVID-19 lockdowns), as shown in Fig. 1. Over this period, age criteria also changed due to increases in the upper age limit for donation, while remaining within the 18–74-year range (Supplementary Table S1).

**Fig. 1:**
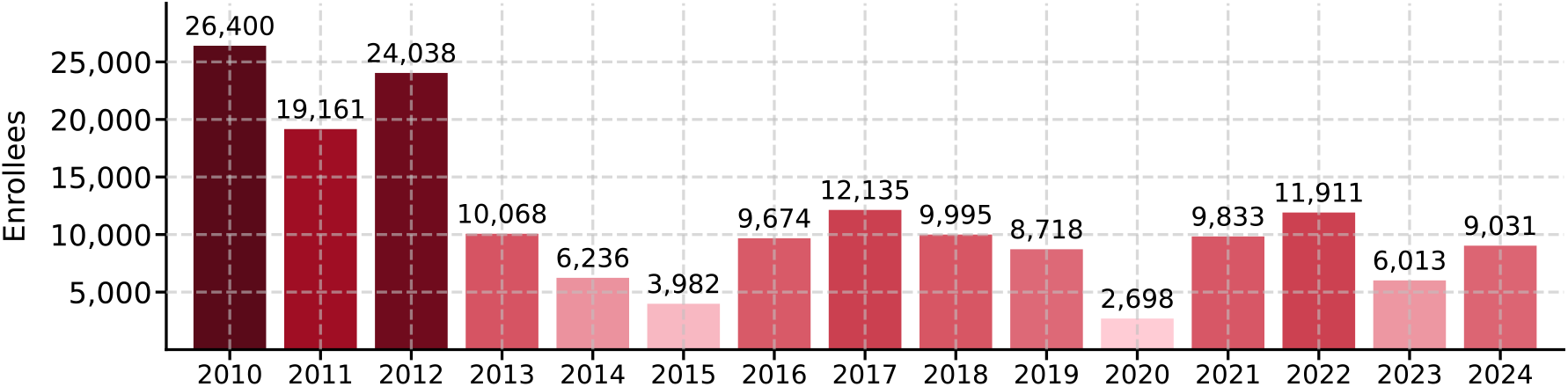
Annual DBDS enrolment in the study population. Number of individuals enrolled in DBDS per calendar year in the study period among those with complete register-based auxiliary variables available as of 31 December of the preceding year; numbers above bars indicate counts.

### Two-stage selection modelling and weighting

To reflect DBDS’s nested two-stage recruitment process, we estimated propensity scores for (i) donation activity and (ii) DBDS enrolment conditional on donation activity; the inverse of their product formed the basis of the inclusion weights. Among DBDS participants, the two stage-specific propensity score distributions differed markedly (Fig. 2a). Stage 1 propensity scores showed a strong mode at low but non-zero values with a rapid decline thereafter and a comparatively short upper tail, whereas stage 2 propensity scores were broader and right-shifted, with an apparent bimodal pattern plausibly reflecting higher enrolment probabilities in the earliest recruitment years, and a more pronounced tail. Stage-specific univariable associations for a prespecified subset of auxiliary variables are shown in Supplementary Fig. S1 and indicate stronger overall selection into donation activity than into DBDS enrolment conditional on donation activity.

**Fig. 2:**
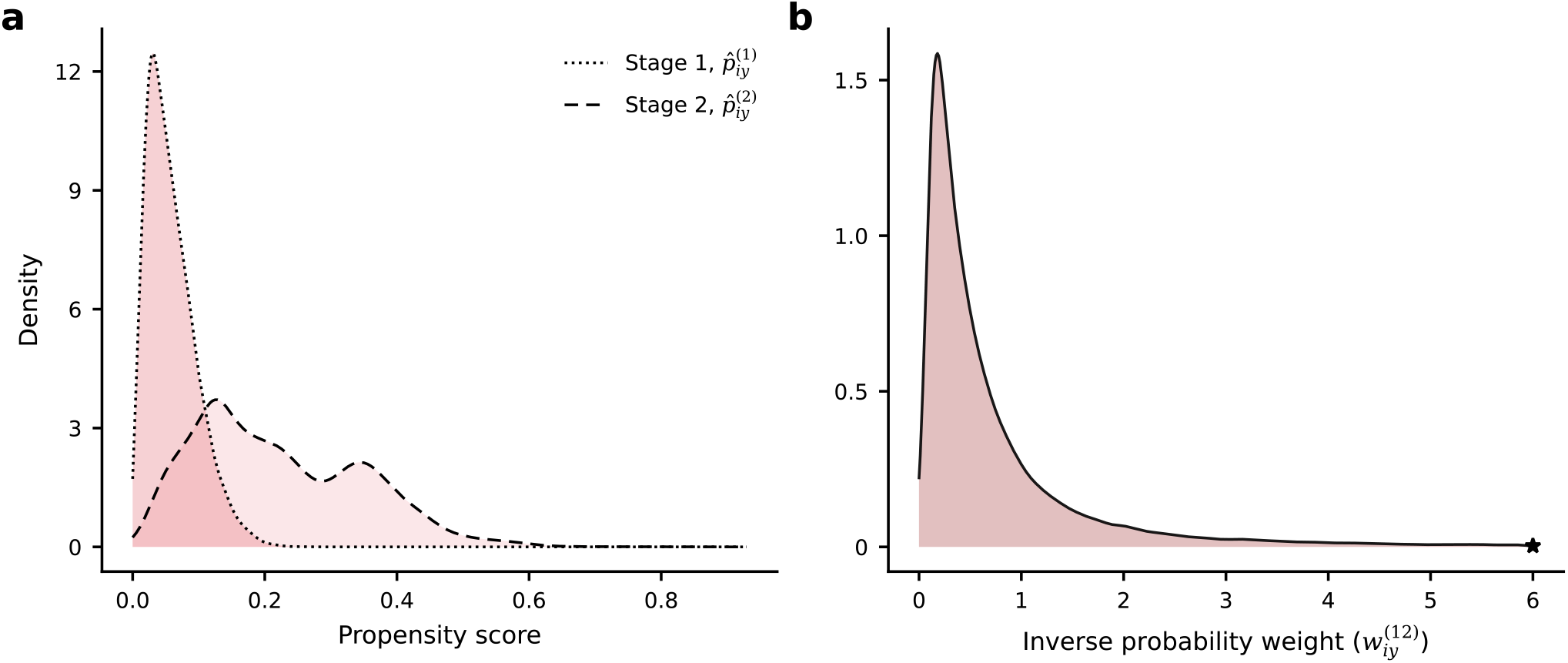
Distributions of stage-specific propensity scores and final inclusion weights. **a**, Probability density functions of the estimated stage-1 and stage-2 propensity scores among DBDS participants. **b**, Probability density function of the final calibrated inclusion weights; the x-axis is truncated, and the star indicates the maximum observed weight in the plotted range.

After stabilization of each stage, winsorization of the combined weights, and subsequent calibration with rescaling to mean 1, the final inclusion weights were right-skewed but generally moderate (Fig. 2b), yielding an effective sample size after IP-weighting of *n*_eff_ = 30,627 (18.03% of 169,893 participants).

### Performance of inclusion weights on auxiliary variables

Next, we quantified the reduction in selection by comparing unweighted and weighted standardized regression coefficients (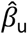 and 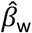) from univariable logistic regression models predicting DBDS inclusion in the target population. Across all main-effect auxiliary variables, the median attenuation was 0.976 (interquartile range (IQR): 0.753–0.997), with near-complete attenuation for variables included among the calibration margins. Sign changes were rare and occurred only when the post-weighting coefficients were near zero.

Fig. 3 shows the unweighted and weighted coefficients for a prespecified subset of auxiliary variables (median attenuation: 0.931; IQR: 0.697–0.997). Strong predictors of inclusion included labour-market detachment (e.g., outside labour force; 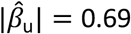), migration background (e.g., immigrant status; 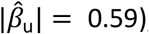, and health-care utilization measures (e.g., unique prescription and diagnosis counts; 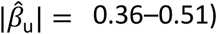. After weighting, most coefficients were attenuated toward zero and residual associations were most evident for selected migration-background and health-care utilization measures, for which 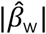 in some cases remained around 0.1–0.2.

**Fig. 3:**
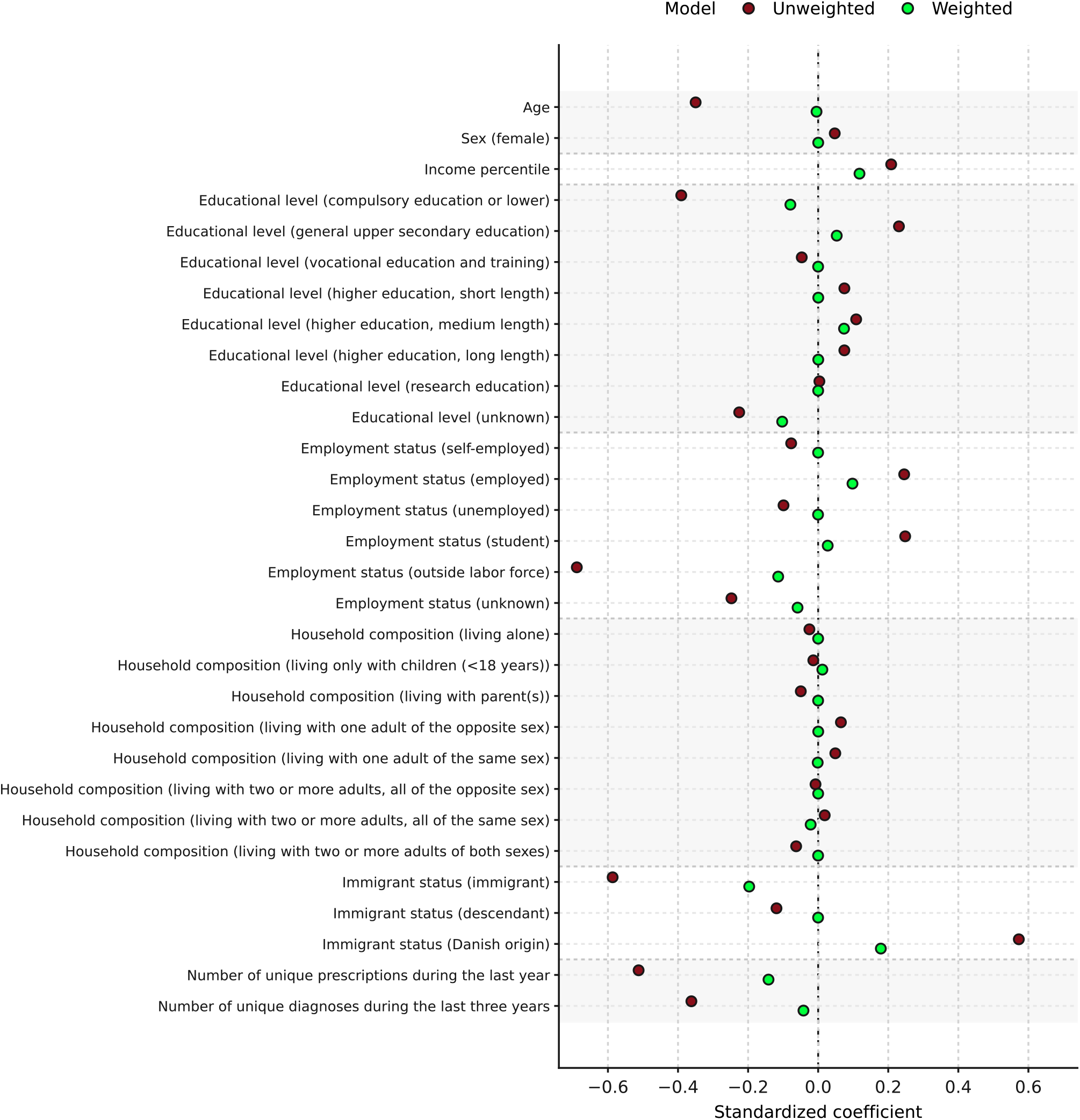
Associations between auxiliary variables and DBDS inclusion before and after weighting. Standardized coefficients for selected auxiliary variables from unweighted and IP-weighted univariable logistic regression models predicting DBDS inclusion. 95% CIs were estimated but are omitted because they are negligible at the plotted scale.

We summarized mean (SD) values for the same prespecified subset of auxiliary variables in the target population, the unweighted DBDS cohort, and the weighted DBDS cohort (Table 1). Unweighted and weighted DBDS estimates are computed on the same analytic set of DBDS participants (n=169,893). In unweighted comparisons, DBDS participants differed systematically from the target population: mean age was lower (38.02 vs 43.25 years), the proportion of females was slightly higher (0.521 vs 0.497), and socioeconomic indicators were shifted upward (e.g., income percentile 57.76 vs 51.92). DBDS participants also had lower health-care utilization in the years preceding enrolment (e.g., unique prescriptions 1.64 vs 2.85; diagnoses 2.30 vs 3.55).

**Table 1:** Register-based characteristics of the target population and DBDS before and after weighting.

| Variable, mean (SD) | Target population | Unweighted DBDS | Weighted DBDS | Change (%) |
| --- | --- | --- | --- | --- |
| Age (years) | 43.251 (15.103) | 38.020 (13.138) | 43.195 (15.002) | + 14 % |
| Sex (female) | 0.497 (0.500) | 0.521 (0.500) | 0.497 (0.500) | - 5 % |
| Income percentile | 51.916 (28.746) | 57.757 (29.177) | 55.243 (28.702) | - 4 % |
| **Highest attained education** |  |  |  |  |
| Compulsory education or lower | 0.245 (0.430) | 0.112 (0.315) | 0.211 (0.408) | + 89 % |
| General upper secondary education | 0.105 (0.307) | 0.190 (0.393) | 0.121 (0.326) | - 36 % |
| Vocational education and training | 0.312 (0.463) | 0.291 (0.454) | 0.312 (0.463) | + 7 % |
| Higher education, short length | 0.049 (0.215) | 0.066 (0.249) | 0.049 (0.215) | - 27 % |
| Higher education, medium length | 0.171 (0.377) | 0.215 (0.411) | 0.200 (0.400) | - 7 % |
| Higher education, long length | 0.091 (0.288) | 0.115 (0.319) | 0.091 (0.288) | - 21 % |
| Research education | 0.008 (0.088) | 0.008 (0.090) | 0.008 (0.088) | - 4 % |
| Unknown | 0.019 (0.137) | 0.003 (0.054) | 0.008 (0.090) | + 183 % |
| **Employment status** |  |  |  |  |
| Self-employed | 0.041 (0.198) | 0.028 (0.164) | 0.041 (0.198) | + 48 % |
| Employed | 0.600 (0.490) | 0.713 (0.452) | 0.647 (0.478) | - 9 % |
| Unemployed | 0.028 (0.165) | 0.015 (0.121) | 0.028 (0.165) | + 87 % |
| Student | 0.114 (0.318) | 0.207 (0.405) | 0.122 (0.327) | - 41 % |
| Outside labour force | 0.176 (0.381) | 0.027 (0.161) | 0.132 (0.339) | + 395 % |
| Unknown | 0.041 (0.198) | 0.010 (0.101) | 0.030 (0.170) | + 190 % |
| **Household composition (evaluated sequentially)** |  |  |  |  |
| Living alone | 0.185 (0.388) | 0.175 (0.380) | 0.185 (0.388) | + 6 % |
| Living only with children (<18 years) | 0.032 (0.177) | 0.030 (0.171) | 0.035 (0.183) | + 15 % |
| Living with parent(s) | 0.093 (0.290) | 0.080 (0.271) | 0.093 (0.290) | + 16 % |
| Living with one adult of the opposite sex | 0.500 (0.500) | 0.533 (0.499) | 0.500 (0.500) | - 6 % |
| Living with one adult of the same sex | 0.034 (0.180) | 0.043 (0.204) | 0.033 (0.179) | - 23 % |
| Living with two or more adults, all the opposite sex | 0.049 (0.217) | 0.048 (0.213) | 0.049 (0.217) | + 3 % |
| Living with two or more adults, all the same sex | 0.010 (0.100) | 0.012 (0.110) | 0.008 (0.090) | - 33 % |
| Living with two or more adults of both sexes | 0.097 (0.296) | 0.080 (0.271) | 0.097 (0.296) | + 22 % |
| **Immigrant status** |  |  |  |  |
| Immigrant | 0.126 (0.332) | 0.019 (0.136) | 0.068 (0.252) | + 263 % |
| Descendant | 0.019 (0.138) | 0.008 (0.087) | 0.019 (0.138) | + 151 % |
| Danish origin | 0.855 (0.352) | 0.973 (0.161) | 0.912 (0.283) | - 6 % |
| Number of unique prescriptions during the last year | 2.853 (3.440) | 1.636 (1.904) | 2.391 (2.685) | + 46 % |
| Number of unique diagnoses during the last three years | 3.545 (4.848) | 2.295 (3.354) | 3.342 (4.389) | + 46 % |

Values are means (SDs) for selected auxiliary variables in the target population, the unweighted DBDS cohort, and the IP-weighted DBDS cohort. Binary variables were coded 0/1; for these variables, the mean corresponds to the proportion. Change (%) is the percent change from unweighted to weighted DBDS means, $(\bar{x}_w - \bar{x}_u)/\bar{x}_u \times 100$ .

After IP-weighting, most variables moved toward target-population values. Age and sex closely matched population values, and differences in socioeconomic and labour-market indicators were markedly reduced. Residual differences were most evident for migration background and health-care utilization, which improved but did not fully reach population levels (e.g., prescriptions 2.39 vs 2.85; diagnoses 3.34 vs 3.55).

### Validation using held-out prescription phenotypes

To assess whether the inclusion weights generalized beyond the auxiliary-variable set used to estimate and calibrate them, we compared prescription prevalences in the weighted DBDS cohort with those in the target population for more granular ATC phenotypes that were not directly included in the propensity models, which used only first-level ATC groups. Specifically, we evaluated 3-character ATC groups defined by ≥1 redeemed prescription in the 1-year window ending on the risk-set date (31 December) and restricted the analysis to groups with ≥500 DBDS participants (42 ATC groups; listed in Supplementary Table S4).

As shown in Fig. 4, IP-weighting shifted DBDS prevalences toward target-population prevalences for most ATC groups, although residual differences remained for a subset of groups, particularly among those reflecting chronic morbidity and sustained treatment use. Several of the larger residual discrepancies were observed in cardiovascular-related therapeutic groups (ATC group C), consistent with persistent health selection in a blood-donor cohort. Across the 42 evaluated groups, the median per-group reduction in absolute log prevalence ratios was 69.7%. Overall, 34 of 42 (81%) ATC groups moved closer to the target-population prevalence after weighting.

**Fig. 4:**
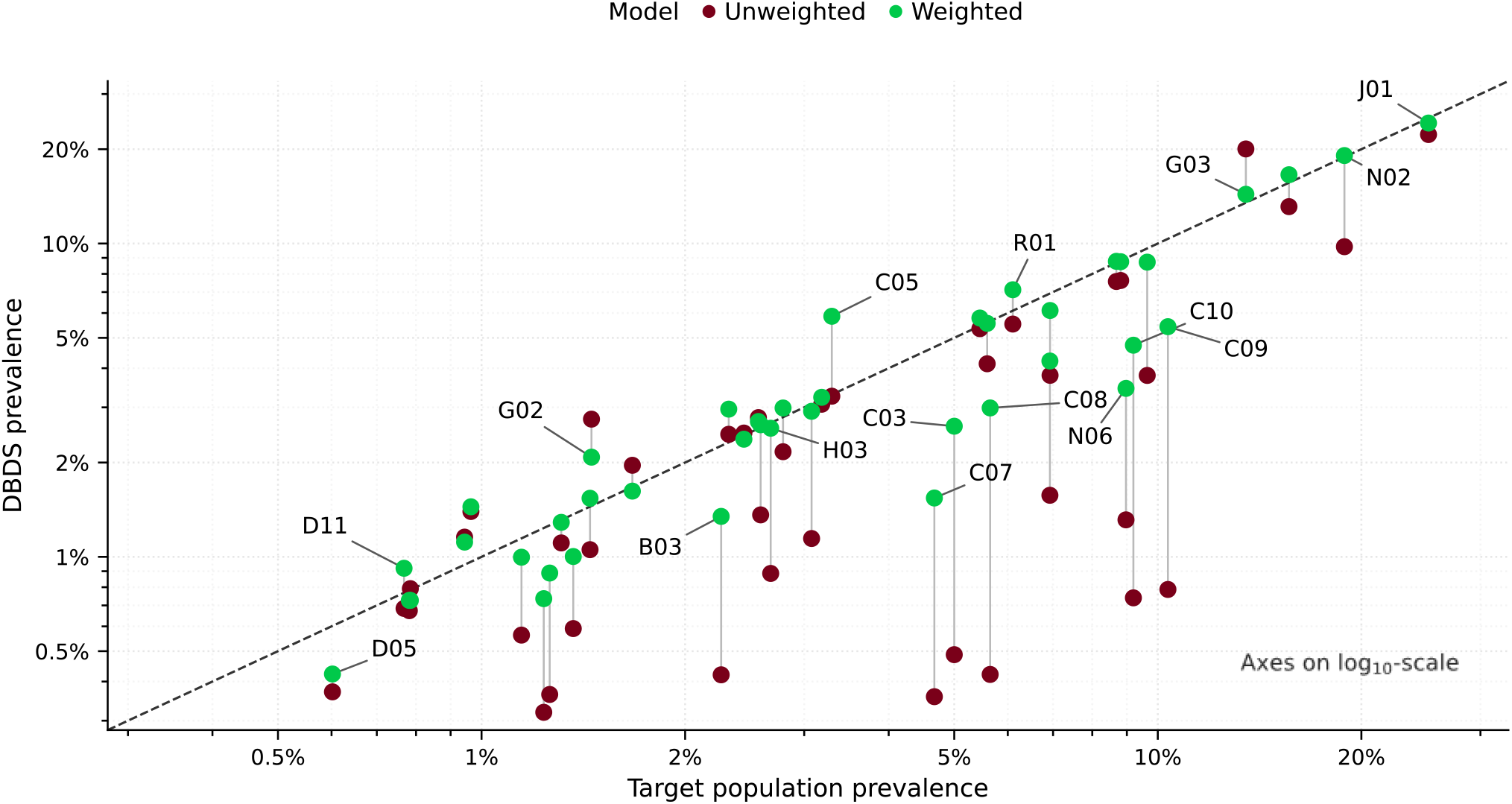
Validation of inclusion weights using held-out prescription phenotypes. Target-population prevalences of 3-character ATC groups compared with unweighted and IP-weighted DBDS prevalences, with ATC-group use defined as ≥1 redemption in the 1-year window ending on the risk-set date (31 December) and restricted to groups with ≥500 DBDS participants. Axes are shown on log_10_-scales; the dashed diagonal indicates equality between DBDS and target-population prevalences. Labels are shown for selected ATC groups to illustrate residual discrepancies across the prevalence range; all evaluated groups are listed in Supplementary Table S4.

### Extension to questionnaire-response weighting

Finally, we extended the two-stage inclusion weights with questionnaire-specific response weights for the first five recurring DBDS questionnaires administered at donation visits (DBDS1–DBDS5). Field periods and respondent counts for DBDS1–DBDS5 are summarized in Supplementary Table S5 and Supplementary Fig. S2, and complete question- and response-option-level results (unweighted and weighted), covering >500 questions and >2,000 response options across DBDS1– DBDS5, are provided in the questionnaire-specific supplementary spreadsheets (Supplementary Data).

Fig. 5 summarizes, for each questionnaire, the eight items with the largest absolute changes in mean after applying the three-stage weights; it is intended to highlight where weighting had the greatest impact within each questionnaire rather than to assess temporal patterns across questionnaire waves. Across DBDS1– DBDS5, the largest changes most often involved poorer self-rated health, physical limitations, sleep/fatigue, selected symptom-severity items, and some health behaviours. Notable examples were poorer self-rated health in DBDS1 (+46.6%), low energy (+36.0%), severe restless-legs discomfort (+33.1%), doctor-diagnosed depression (+31.0%), and daily/almost daily wine drinking (+23.4%) in DBDS2, back pain due to herniated disc (+27.3%), fertility problems (+25.8%), and severe menstrual pain (+25.6%) in DBDS3, limitations in climbing several flights of stairs in DBDS4 (+43.2%), and nocturnal urination together with sleep-, attention-, depression-, and stress-related items in DBDS5 (approximately +23.8% to +25.5%). A smaller number of items moved in the opposite direction, most notably non-cigarette tobacco intensity in DBDS1 (cheroots/cigars per day −22.1%; pipefuls per day −19.7%) and daytime urinary leakage at ages 6–7 in DBDS5 (−21.2%).

**Fig. 5:**
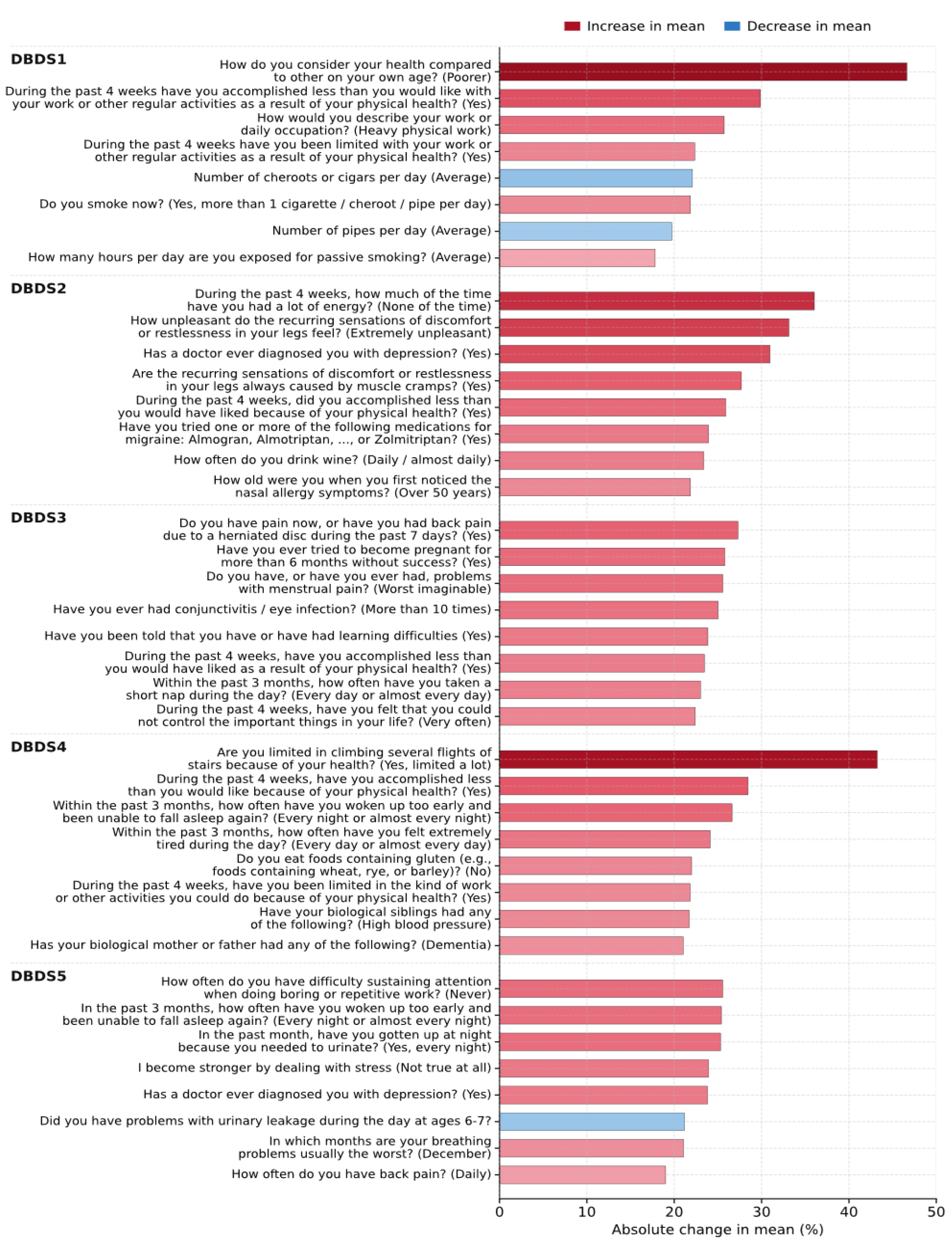
Changes in questionnaire-based means after weighting. Absolute percent changes in mean response values after three-stage IP-weighting, 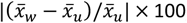, shown for the eight eligible items with the largest absolute change within each DBDS1–DBDS5 questionnaire. Categorical questions were eligible if each non-’don’t know’ response option had ≥500 respondents; continuous questions were eligible if they had ≥500 respondents. Red bars indicate increases in mean response values and blue bars indicate decreases.

## Discussion

Selective participation is increasingly recognized as a challenge to external validity in large volunteer-based cohorts and biobanks, with recent studies showing that IP-weighting can reduce selection on observed characteristics when suitable external benchmarks exist^8–10^. However, applications have largely focused on settings in which study participation is modelled as a single selection process, rather than across multiple stages at recruitment or during subsequent participation. In the present work, we developed and applied a multi-stage IP-weighting framework to DBDS, a nationwide biobank in which inclusion is shaped by two stages of selection—having an eligible donation visit and subsequently enrolling in DBDS—and may be followed by additional stages of participation such as questionnaire response.

In the unweighted DBDS cohort, participants differed systematically from the target population across several demographic, socioeconomic, and health-related characteristics. After applying the two-stage inclusion weights, the DBDS cohort moved substantially closer to target-population values across most evaluated variables, including close alignment for core demographic and socioeconomic measures. This was also evident in the predictive evaluation: standardized coefficients from univariable inclusion models were markedly attenuated towards zero after weighting, indicating substantially reduced selection on the observed register-based auxiliary variables. Validation using held-out prescription-based phenotypes likewise showed improved alignment with target-population prevalences after weighting. Extending the framework with a third, questionnaire-response stage further showed that the same staged logic can be carried forward to downstream participation processes after enrolment.

In comparison with recent IP-weighting efforts in UKB^8–10^, our findings are qualitatively consistent with the notion that high-dimensional auxiliary adjustment can substantially reduce selection on observed variables. At the same time, UKB applications have largely focused on settings in which study participation is modelled as a single selection process, despite UKB also having later participation opportunities such as repeat assessments, imaging, and web-based questionnaires^6,7^. More generally, however, a staged formulation may be preferable whenever selection operates over more than one stage, whether within recruitment itself or through additional stages after enrolment. This is particularly relevant in DBDS, where enrolment is preceded by entry into a health-screened donor system—an additional, health-linked filter that UKB did not impose. Moreover, whereas UKB recruitment was concentrated within a fixed five-year period, DBDS recruitment has continued since 2010, so enrolment may vary more across calendar years, motivating estimation on annual inclusion-anchored risk sets. An additional practical distinction is that the DBDS framework leverages nationwide administrative registers with near-complete coverage and yearly alignment to those risk sets, whereas UKB weighting studies have relied more heavily on surveys or census-derived benchmarks. This strengthens coverage of demographic, socioeconomic, and healthcare-related variables and allows risk sets and auxiliary variables to be defined consistently over time, although registers capture self-reported behaviours, symptom burden, and related determinants of selection less directly.

For DBDS inclusion, the two-stage formulation is not only conceptually appropriate but also practically valuable. It separates selection into donation from selection into DBDS enrolment, allows each stage to be modelled on its natural risk set, and can reveal selection patterns that a single direct model of DBDS inclusion from the target population would tend to mask. In our data, observed associations were generally stronger at the donation stage than at the DBDS enrolment stage conditional on donation, indicating that the largest observable selection occurred in the transition from the annual population risk sets into the active donor pool. A related limitation, however, is that stage-2 probabilities may be estimated less precisely in calendar years with lower DBDS enrolment, including periods affected by temporary interruptions such as the COVID-19 lockdowns.

A central implication of the results is that some characteristics are inherently difficult to generalize from DBDS to the target population, even with extensive register-based adjustment, because selection involves structural barriers and limited overlap with parts of that population. Blood donors constitute a highly selected subset of the population because of donor screening and self-selection, a phenomenon widely described as the healthy donor effect^13,25–28^. This selection mechanism is expected to be strongest for morbidity and healthcare utilization, precisely the domains in which donor eligibility criteria, deferrals, and health-related self-selection operate most directly. In addition, DBDS enrolment and invitation to donation-visit questionnaires occur in a Danish clinical setting and rely on language proficiency, comprehension, and willingness to participate; participation may therefore remain systematically lower among groups facing language-related barriers or differential institutional contact. In our analyses, this was reflected in persistent residual discrepancies for migration background and healthcare utilization, consistent with limited overlap in parts of the target population and with the fact that these variables were not fully enforced as calibration margins in the final raking specification.

More generally, when parts of the target population have near-zero probability of entering the donation- and-enrolment pipeline, or of subsequently contributing downstream data such as questionnaire responses, weighting can reduce but not eliminate differences without inducing extreme weights or relying on strong extrapolation^19,29,30^. A staged formulation does not remove these structural constraints, but it can help localize them by clarifying whether limited overlap arises primarily at the donation stage, the subsequent enrolment stage, or later participation stages. Consistent with these constraints, selection on observed auxiliary variables was markedly reduced, but at the cost of a lower effective sample size, reflecting the strong structural filtering inherent to donation-based recruitment.

The questionnaire extension further shows how staged weighting can be used for downstream participation processes that occur after enrolment and are often ignored when study participation is modelled as a single selection process. In DBDS, many key phenotypes, including self-reported behaviours, symptoms, and health history, are collected through repeated questionnaires administered within the donation setting, with response windows and response patterns varying across questionnaire waves. Combining the two-stage inclusion weights with questionnaire-specific response weights therefore provides a coherent way to generalize questionnaire-based outcomes from respondents to the same target population as the primary inclusion weights, while separating enrolment selection from response selection in nested deep-phenotyping settings.

An important next step is to extend the same framework to DBDS biomarker panels measured in stored plasma samples and to genetic analyses based on the DBDS genomic resource^15,16^, to quantify how reweighting changes biomarker distributions and downstream associations, paralleling what has been documented for UKB. For genetics-focused applications, it may also be preferable to define the target population more closely to match the intended analysis population—for example, by restricting the target when analyses are limited to specific ancestry groups—to improve overlap and weight stability. More broadly, the framework should be applicable beyond DBDS to other cohorts and biobanks in which selection unfolds over more than one stage, whether through eligibility filtering, repeat-contact participation, or nested questionnaire and biomarker collection.

Overall, our findings support multi-stage IP-weighting as a feasible strategy for improving the external validity of DBDS-based analyses, particularly for many demographic and socioeconomic characteristics, while also clarifying where structural features of blood-donor recruitment impose fundamental limits on representativeness.

## Data availability

Person-level data from DBDS needed to reproduce this study cannot be made publicly available due to confidentiality legislation. Furthermore, this study is based on pseudonymized register data located on a secure platform at Statistics Denmark. Meta-data and programs are available from the authors upon reasonable request and with permission of the DBDS steering committee, the Ethical Committee, and the Danish Data Protection Agency. Enquiries about legal possibilities for accessing these data within DBDS, scripts/codes and further information should be addressed to the corresponding author.

## Statements and declarations

## Acknowledgements

We thank the Danish blood donors for their valuable participation in the Danish Blood Donor Study, and the staff at the blood centres for making this study possible.

## Ethics approval

DBDS was approved by the Central Denmark (1–10-72–95-13) and Zealand (SJ-740) Regional Committees on Health Research Ethics and the Data Protection Agency (P-2019–99). Oral and written informed consent was obtained from all DBDS participants. All methods were carried out in accordance with the relevant guidelines and regulations (including the Declaration of Helsinki).

## Author contributions

M.D. proposed the initial study idea, J.T. developed the analytical framework, and S.R.O. provided overall supervision and scientific guidance throughout the study. J.T. curated the data, performed the analyses, and drafted the manuscript. J.T., M.D., D.H., L.A.N.C., K.M.D., B.M., and S.R.O. interpreted the results. E.S., C.E., O.B.V.P., and S.R.O. contributed to cohort oversight and research infrastructure. J.D., C.M., L.H., L.J.E.Q., M.T.B., B.A., T.F.H., H.H., K.R., E.S., C.E., O.B.V.P., T.H., and A.J.S. reviewed and commented on the manuscript. All authors approved the final version before submission.

