## Supplementary information for "Multi-stage reweighting to correct for participation bias in a nationwide biobank with nested recruitment"

**Address for correspondence:**

### AGE ELIGIBILITY AND SAMPLING-YEAR RISK SETS

For each sampling year, the annual population risk set comprised Danish residents who had at least one day during that calendar year meeting the DBDS age-eligibility criteria and who had not enrolled in DBDS before that year. DBDS recruitment commenced on 1 March 2010 (so the 2010 sampling frame covered 1 March–31 December). Age limits changed over calendar time:

**Supplementary Table S1**. Eligible age range for DBDS participation by sampling year.

| Sampling year(s) | Minimum age | Maximum age |
|---|---|---|
| 2010–2018 | 18 years | <67 years |
| 2019–2020 | 18 years | <70 years |
| 2021 | 18 years | <70 years (to 31 Oct);<br><75 years (from 1 Nov) |
| 2022 onward | 18 years | <75 years |

The upper age thresholds used for constructing the sampling frame followed the eligibility rules for continuing donors. This choice avoids conditioning risk-set membership on prior donation history: using separate limits for new and continuing donors would either require excluding DBDS participants who enrolled above the new-donor limit or, if retained, would imply that the oldest age strata in the risk sets are necessarily blood donors (i.e., introducing donation-history information into the sampling frame).

### AUXILIARY VARIABLES

Auxiliary variables were obtained from Danish national administrative registers and defined consistently for all individuals within each sampling-year risk set. Unless otherwise stated below, variables were anchored to the relevant sampling year. Medication history was summarized over the previous 12 months, and hospital diagnoses were summarized over the previous 3 years.

**Propensity-model coding**. All multi-category variables were expanded to binary indicator columns (one per category) using a reference category. Variables that were already binary were entered directly. Age and income percentile were represented using natural spline bases with 4 and 3 degrees of freedom, respectively. For the prescription and diagnosis count variables, we used a two-part parameterization consisting of an indicator for any history (count > 0) and log(1 + count). DBDS sampling year was included as a design variable and represented by one indicator column per year.

**Supplementary Table S2**. Auxiliary variables and coding used in the propensity models.

| Variable | Definition / categories | Coding in propensity models |
|---|---|---|
| (Legal) sex | Male; Female. | Entered directly (binary). |
| Age | Age in years (continuous). | Natural spline basis, 4 df. |
| Civil status | Unmarried; Married; Divorced; Widowed. | Binary indicator columns. |
| Household size | 1; 2; 3; 4; 5; ≥6 persons. | Binary indicator columns. |
| Household composition (evaluated sequentially) | Living alone; Living only with children (<18 years); Living with parent(s); Living with one adult of the opposite sex; Living with one adult of the same sex; Living with two or more adults all the opposite sex; Living with two or | Binary indicator columns. |

| | | |
|---|---|---|
| | more adults all the same sex; Living with two or more adults of both sexes. | |
| Number of children | 0; 1; 2; 3; 4; ≥5 children. | Binary indicator columns. |
| Region of residence | Central Denmark Region; Capital Region of Denmark; North Denmark Region; Region of Southern Denmark; Region Zealand. | Binary indicator columns. |
| Country of origin | Danish origin; Other western countries; MENAPT countries; Rest of the world. | Binary indicator columns. |
| Immigrant status | Danish origin; Immigrant; Descendant. | Binary indicator columns. |
| Highest attained education | Compulsory education or lower; General upper secondary education; Vocational education and training; Higher education (short); Higher education (medium); Higher education (long); Research education; Unknown. | Binary indicator columns. |
| Employment status | Self-employed; Employed; Unemployed; Student; Outside labour force; Unknown. | Binary indicator columns. |
| Profession | Agriculture, forestry, and fishing; Business services; Construction; Culture, leisure, other services; Finance and insurance; Industry, raw materials, utilities; Information and communication; Public administration, education, and healthcare; Real estate and leasing; Trade and transport; Unknown. | Binary indicator columns. |
| Income percentile | Percentile rank (1-100) within the Danish population aged 17-74 years on 31 December of the year preceding the sampling year (continuous). | Natural spline basis, 3 df. |
| Unique redeemed prescriptions (previous 12 months) | Count of distinct redeemed prescription drug codes during the previous 12 months (continuous). | Indicator I(count>0) and log(1+count). |
| Unique hospital diagnoses (previous 3 years) | Count of distinct hospital diagnosis codes during the previous 3 years (continuous). | Indicator I(count>0) and log(1+count). |
| Charlson comorbidity index | Non-age-adjusted Charlson comorbidity index categorized as | Binary indicator columns. |

| | | |
|---|---|---|
| | 0; 1; 2+ (based on diagnoses during the previous 3 years). | |
| First-level ATC groups | Binary indicators for each first-level ATC group (any redemption in the previous 12 months). | Entered directly (binary indicators). |
| First-level ICD-10 chapters | Binary indicators for each first-level ICD-10 chapter (any hospital diagnosis in the previous 3 years). | Entered directly (binary indicators). |
| Sampling year | Calendar year defining the sampling frame (2010–2024) | Binary indicator columns (one per year). |

<u>Notes on selected auxiliary variables</u>

**Country of origin and immigrant status:** These variables followed Statistics Denmark's register-based definitions, which combine information on the individual's and parents' country of birth and citizenship. Danish origin indicates that at least one parent was born in Denmark and held a Danish citizenship. Immigrant denotes individuals born abroad with no parent meeting the above requirements, whereas descendant denotes persons born in Denmark with no parent meeting the above requirements; when parental information was unavailable, classification followed Statistics Denmark conventions based on the individual's country of birth and citizenship. Immigrant status captures generational position (Danish origin, immigrant, descendant), whereas country of origin captures broad geographic origin. For modelling, country of origin was grouped as Danish origin, other Western countries, MENAPT (Middle East, North Africa, Pakistan and Turkey) countries, and the rest of the world. Western/non-Western followed Statistics Denmark's classification, and MENAPT corresponds to the grouping used in Denmark's national Integration Barometer.

**Household composition:** Derived from co-resident information and categorized as living alone, living only with children (<18 years), living with parent(s), or adult cohabitation structures defined by the number and sex of co-resident adults. The variable was constructed using a sequential rule set (living alone -> living only with children -> living with parent(s) -> adult cohabitation categories) so that each person was assigned exactly one, mutually exclusive category.

**Highest attained education:** Categories reflect Danish education-register groupings. Compulsory education corresponds to primary and lower secondary schooling. Higher education was grouped by typical program length: short-cycle (about 2 years), medium-cycle (about 3–4 years, e.g. bachelor's level), and long-cycle (about 5–6 years, e.g. master's-level). Research education includes doctoral-level qualifications (e.g. PhD). 'Unknown' denotes missing or unclassifiable information.

**Medication and hospital history:** Counts of unique redeemed prescriptions and unique hospital diagnoses reflect the number of distinct ATC codes redeemed during the prior 12 months and distinct ICD-10 diagnosis codes recorded during the prior 3 years, respectively. We used a 3-year diagnosis look-back (rather than 1 year) to better capture chronic conditions and reduce sensitivity to short-term events. First-level ATC groups and ICD-10 chapters were included as binary indicators of any redemption/diagnosis within each broad category, complementing the code-level counts.

**Charlson comorbidity index:** A non-age-adjusted Charlson score was computed from hospital diagnoses in the 3-year look-back window and categorized as 0, 1, or 2+. Values above 1 were uncommon among DBDS participants, consistent with blood-donor eligibility, and were therefore pooled. The index was kept non-age-adjusted because age was included separately in the propensity models and all two-way interactions were considered, allowing flexible joint effects of age and comorbidity.

### CALIBRATION TARGETS (RAKING MARGINS)

We calibrated the combined stage-1+2 inclusion weights by iterative proportional fitting (raking) to selected margins computed in the sampling-year risk sets (Supplementary Table S3). Age, income percentile and sampling year were coarsened to avoid sparse cells. Starting from base margins (sex-by-age group and sampling-year group), additional main-effect margins were selected using a forward-selection procedure that prioritized improvements in overall covariate balance while monitoring effective sample size (ESS).

At each step $t$, we evaluated candidate margins one at a time by calibrating the weights and quantifying improvement in balance across all main-effect auxiliary variables. Balance was measured using standardized mean differences (SMDs) between DBDS and the target population:

$$\mathrm{SMD}_j = \frac{\bar{x}_{\mathrm{DBDS},j} - \bar{x}_{\mathrm{Target},j}}{\sqrt{\left(s^2_{\mathrm{DBDS},j} + s^2_{\mathrm{Target},j}\right)/2}}.$$

Let $\mathrm{SMD}_{u,j}$ denote the unweighted SMD for variable $j$ and $\mathrm{SMD}^c_{w,j}$ the SMD after calibration with candidate margin $c$ (computed using weighted DBDS means and variances). We defined the proportional reduction in absolute SMD as $r_j(c) = 1 - \left|\mathrm{SMD}^c_{w,j}\right| / \left|\mathrm{SMD}_{u,j}\right|$, and summarized overall improvement for candidate $c$ as the median across auxiliary variables, $R(c) = \mathrm{median}_j\, r_j(c)$. Effective sample size (ESS) of the calibrated weights was computed as $\mathrm{ESS}(c) = \left(\sum_i w_i\,(c)\right)^2 / \sum_i w_i\,(c)^2$.

To account for potential loss of precision from additional calibration constraints, we defined the relative ESS loss at step t as: $\Delta E(c) = (\mathrm{ESS}_t - \mathrm{ESS}(c))/\mathrm{ESS}_t$. We then ranked candidate margins using a gain-per-ESS-loss score:

$$U(c) = \frac{R(c) - R_t}{\max(\Delta E(c), \varepsilon)}.$$

At each step we selected the candidate margin c* that maximized U(c) among candidates with a minimum improvement $R(c) - R_t \geq \delta$, using δ = 0.005. The forward-selection procedure stopped when no remaining candidate achieved an improvement of at least δ. A small constant ε was used to avoid division by values close to zero. In case of $\Delta E(c) < 0$, we picked the candidate that maximized $R(c) - R_t \geq \delta$.

**Supplementary Table S3**. Calibration targets used for raking of the combined inclusion weights.

| Calibration target | Levels / definition | Source variable (or grouping) |
|---|---|---|
| Sex and age | Female (binary indicator) * (17–24; 25–34; 35–44; 45–54; 55–64; 65–74 years). | Interaction: (legal) sex * age (grouped) |
| Civil status | Widow(er) (binary indicator). | Civil status |
| Household size | Household sizes 1, 2, 3, 4, 5, and 6 (binary indicators). | Household size |
| Household composition | Living alone; living with parent(s); living with one adult of the opposite sex; living with ≥2 adults (all opposite sex); living with ≥2 adults (both sexes) (binary indicators). | Household composition |

| | | |
|---|---|---|
| Number of children | Number of children: 4 and 5+ (binary indicators). | Number of children |
| Region of residence | North Denmark Region and Capital Region of Denmark (binary indicators). | Region of residence |
| Migration background | Descendant (binary indicator). | Immigrant status |
| Education | Vocational education; short-cycle higher education; long-cycle higher education; research education (binary indicators). | Highest attained education |
| Employment | Self-employed (binary indicator). | Employment status |
| Income | 61–80 (top quintile) only (binary indicator). | Income percentile grouped into quintiles (1–20, 21–40, 41–60, 61–80, 81–100) |
| Profession | Agriculture/forestry/fishing; finance/insurance; real estate/leasing (binary indicators). | Profession |
| Comorbidity | Charlson comorbidity index = 1 (binary indicator). | Charlson comorbidity index |
| Medication history | First-level ATC group D, H, S and V (binary indicators: any redemption in prior year). | ATC groups |
| Hospital history | First-level ICD-10 chapter indicators: 09, 10, 12, 14, 15, 16, 17, 20 (binary indicators; 3-year lookback). | ICD-10 chapters |
| Sampling year group | 2010; 2011; 2012; 2013–2014; 2015–2016; 2017–2018; 2019–2020; 2021–2022; 2023–2024. | Sampling year (grouped) |

### Stage-specific associations with donation activity and DBDS enrolment

To illustrate how the two recruitment stages differed in their selection patterns, we fitted unweighted univariable logistic regression models for donation activity (stage 1) and DBDS enrolment conditional on donation activity (stage 2) using the same prespecified subset of auxiliary variables shown in the main-text coefficient figure. Supplementary Fig. S1 shows that associations were generally stronger for stage 1 than for stage 2, indicating that the largest observable selection occurred in the transition from the annual population risk sets into the active donor pool, whereas subsequent enrolment into DBDS among donors showed weaker but still patterned selection.

**Supplementary Fig. S1.** Standardized coefficients from unweighted univariable logistic regression models for stage 1 (donation activity in the annual population risk set) and stage 2 (DBDS enrolment conditional on donation activity) for a prespecified subset of auxiliary variables. Coefficients are shown to illustrate how the selection patterns differed between the transition from the annual-population risk set into the active donor pool and the subsequent transition from active donor to DBDS participant.

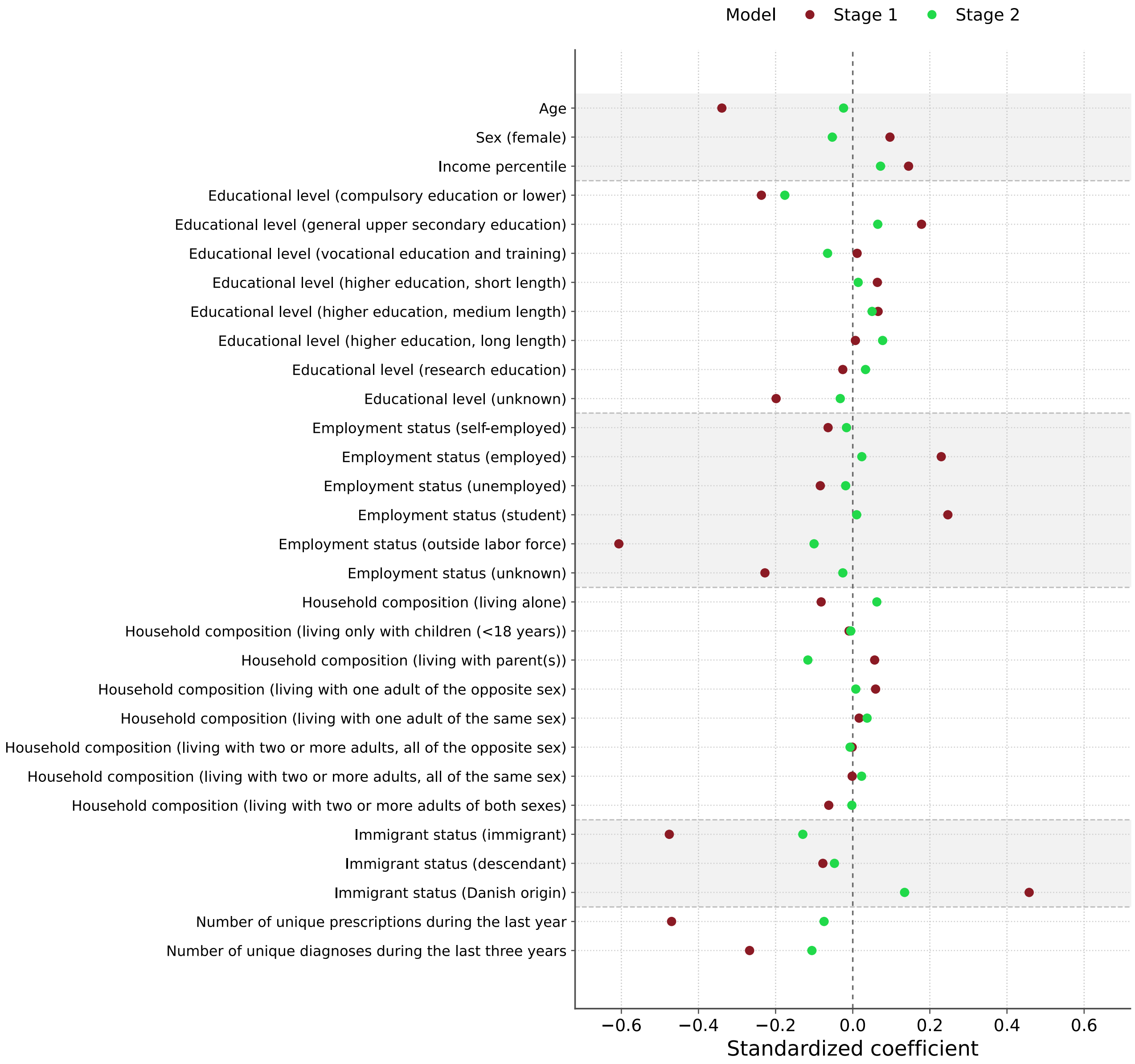


### HELD-OUT PHENOTYPES FOR VALIDATION OF INCLUSION WEIGHTS

To evaluate generalization beyond the exact auxiliary-variable set used for propensity estimation and calibration, we evaluated more granular prescription phenotypes that were not used directly in the propensity models. Specifically, we considered 3-character ATC groups (for example, A06 and C07), whereas the propensity models included only first-level ATC-group indicators (A-V) within the prior 12 months. ATC-group use was defined as ≥1 redeemed prescription in the 1-year window ending on the risk-set date (31 December). We restricted the validation set to 3-character ATC groups with ≥500 DBDS participants (counted in participants' most recent risk set), yielding 42 ATC groups. For each group, we estimated target-population prevalence, unweighted DBDS prevalence, and IP-weighted DBDS prevalence, and computed the absolute log prevalence ratio $|\log(p_{\mathrm{DBDS}}/p_{\mathrm{target}})|$, before and after weighting.

We also explored an analogous validation based on 3-character ICD-10 diagnoses, but representation across ICD-10 chapters among codes meeting the prevalence threshold was highly uneven (including overrepresentation of chapter Z and sparse/no representation in multiple other chapters), limiting interpretability of a chapter-spanning validation set. We therefore used 3-character ATC groups for the primary held-out phenotype validation.

**Supplementary Table S4.** Held-out 3-character ATC groups used for validation of inclusion weights.

| ATC3 code | ATC chapter | ATC 3-character group name | Prevalence (target / DBDS / weighted DBDS) | Absolute log prevalence ratio (DBDS / target) | Absolute log prevalence ratio (weighted DBDS / target) |
|---|---|---|---|---|---|
| A01 | A — Alimentary tract and metabolism | Stomatological preparations | 1.4% / 1.1% / 1.5% | 0.317 | 0.061 |
| A02 | A — Alimentary tract and metabolism | Drugs for acid related disorders | 9.6% / 3.8% / 8.7% | 0.933 | 0.101 |
| A03 | A — Alimentary tract and metabolism | Drugs for functional gastrointestinal disorders | 1.1% / 0.6% / 1.0% | 0.711 | 0.139 |
| A06 | A — Alimentary tract and metabolism | Drugs for constipation | 1.3% / 0.4% / 0.9% | 1.243 | 0.351 |
| B03 | B — Blood and blood forming organs | Antianemic preparations | 2.3% / 0.4% / 1.3% | 1.683 | 0.518 |
| C03 | C — Cardiovascular system | Diuretics | 5.0% / 0.5% / 2.6% | 2.328 | 0.649 |
| C05 | C — Cardiovascular system | Vasoprotectives | 3.3% / 3.3% / 5.9% | 0.012 | 0.575 |
| C07 | C — Cardiovascular system | Beta blocking agents | 4.7% / 0.4% / 1.5% | 2.569 | 1.109 |
| C08 | C — Cardiovascular system | Calcium channel blockers | 5.7% / 0.4% / 3.0% | 2.596 | 0.637 |
| C09 | C — Cardiovascular system | Agents acting on the renin-angiotensin system | 10.4% / 0.8% / 5.4% | 2.577 | 0.646 |
| C10 | C — Cardiovascular system | Lipid modifying agents | 9.2% / 0.7% / 4.7% | 2.521 | 0.664 |
| D01 | D — Dermatologicals | Antifungals for dermatological use | 5.5% / 5.3% / 5.8% | 0.020 | 0.058 |
| D05 | D — Dermatologicals | Antipsoriatics | 0.6% / 0.4% / 0.4% | 0.484 | 0.353 |
| D06 | D — Dermatologicals | Antibiotics and chemotherapeutics | 3.2% / 3.1% / 3.2% | 0.037 | 0.014 |

| | | | | | |
|---|---|---|---|---|---|
| | | for dermatological use | | | |
| D07 | D — Dermatologicals | Corticosteroids, dermatological preparations | 8.8% / 7.6% / 8.7% | 0.145 | 0.007 |
| D10 | D — Dermatologicals | Anti-acne preparations | 1.7% / 2.0% / 1.6% | 0.159 | 0.030 |
| D11 | D — Dermatologicals | Other dermatological preparations | 0.8% / 0.7% / 0.9% | 0.115 | 0.180 |
| G01 | G — Genito urinary system and sex hormones | Gynecological antiinfectives and antiseptics | 0.8% / 0.8% / 0.7% | 0.009 | 0.077 |
| G02 | G — Genito urinary system and sex hormones | Other gynecologicals | 1.5% / 2.8% / 2.1% | 0.638 | 0.358 |
| G03 | G — Genito urinary system and sex hormones | Sex hormones and modulators of the genital system | 13.5% / 20.0% / 14.4% | 0.395 | 0.062 |
| G04 | G — Genito urinary system and sex hormones | Urologicals | 3.1% / 1.1% / 2.9% | 0.989 | 0.054 |
| H02 | H — Systemic hormonal preparations (excl. sex hormones and insulins) | Corticosteroids for systemic use | 2.6% / 1.4% / 2.6% | 0.642 | 0.020 |
| H03 | H — Systemic hormonal preparations (excl. sex hormones and insulins) | Thyroid therapy | 2.7% / 0.9% / 2.6% | 1.106 | 0.039 |
| J01 | J — Antiinfectives for systemic use | Antibacterials for systemic use | 25.1% / 22.3% / 24.2% | 0.121 | 0.036 |
| J02 | J — Antiinfectives for systemic use | Antimycotics for systemic use | 2.6% / 2.8% / 2.7% | 0.081 | 0.052 |
| J05 | J — Antiinfectives for systemic use | Antivirals for systemic use | 2.3% / 2.5% / 3.0% | 0.059 | 0.244 |

| | | | | | |
|---|---|---|---|---|---|
| J07 | J — Antiinfectives for systemic use | Vaccines | 1.0% / 1.4% / 1.4% | 0.370 | 0.404 |
| M01 | M — Musculo-skeletal system | Antiinflammatory and antirheumatic products | 15.6% / 13.1% / 16.6% | 0.176 | 0.059 |
| M03 | M — Musculo-skeletal system | Muscle relaxants | 1.4% / 0.6% / 1.0% | 0.839 | 0.310 |
| N02 | N — Nervous system | Analgesics | 18.9% / 9.8% / 19.1% | 0.659 | 0.011 |
| N05 | N — Nervous system | Psycholeptics | 6.9% / 1.6% / 4.2% | 1.483 | 0.496 |
| N06 | N — Nervous system | Psychoanaleptics | 9.0% / 1.3% / 3.4% | 1.922 | 0.956 |
| N07 | N — Nervous system | Other nervous system drugs | 1.2% / 0.3% / 0.7% | 1.354 | 0.519 |
| P01 | P — Antiparasitic products, insecticides and repellents | Antiprotozoals | 2.4% / 2.5% / 2.4% | 0.017 | 0.029 |
| P02 | P — Antiparasitic products, insecticides and repellents | Anthelmintics | 0.9% / 1.2% / 1.1% | 0.203 | 0.166 |
| R01 | R — Respiratory system | Nasal preparations | 6.1% / 5.5% / 7.1% | 0.098 | 0.154 |
| R03 | R — Respiratory system | Drugs for obstructive airway diseases | 6.9% / 3.8% / 6.1% | 0.602 | 0.125 |
| R05 | R — Respiratory system | Cough and cold preparations | 2.8% / 2.2% / 3.0% | 0.254 | 0.067 |
| R06 | R — Respiratory system | Antihistamines for systemic use | 5.6% / 4.1% / 5.6% | 0.303 | 0.006 |
| S01 | S — Sensory organs | Ophthalmologicals | 8.7% / 7.6% / 8.8% | 0.138 | 0.010 |
| S02 | S — Sensory organs | Otologicals | 1.3% / 1.1% / 1.3% | 0.169 | 0.018 |
| S03 | S — Sensory organs | Ophthalmological and otological preparations | 0.8% / 0.7% / 0.7% | 0.150 | 0.074 |

### DBDS QUESTIONNAIRES (DBDS1–DBDS5)

Across DBDS1–DBDS5, we first excluded responses timestamped before recorded DBDS enrolment within the study period (n=989). We then applied the questionnaire-specific exclusions shown below.

**Supplementary Table S5**. Field periods, analytic respondent counts, and exclusions for DBDS1–DBDS5.

| Questionnaire | Field period | Respondents included | Exclusions |
|---|---|---|---|
| DBDS1 | Mar 2010–May 2015 | 83,137 | No exclusions due to register-variable availability (response at enrolment). |
| DBDS2 | Jun 2015–May 2018 | 52,302 | Excluded 60 responses lacking register variables (31 Dec of preceding year). |
| DBDS3 | Jun 2018–Mar 2020 | 46,278 | Excluded 72 responses lacking register variables (31 Dec of preceding year). |
| DBDS4 | Nov 2020–Mar 2023 | 52,784 | Excluded 83 responses lacking register variables (31 Dec of preceding year). |
| DBDS5 | Mar 2023–May 2025 | 44,286 | Restricted to responses through 31 Dec 2024; excluded 55 responses lacking register variables (31 Dec of preceding year). |

**Supplementary Fig. S2**. Field periods of the five DBDS questionnaires. The dashed vertical line marks the end of the study period (31 Dec 2024); DBDS5 responses after this date were excluded.

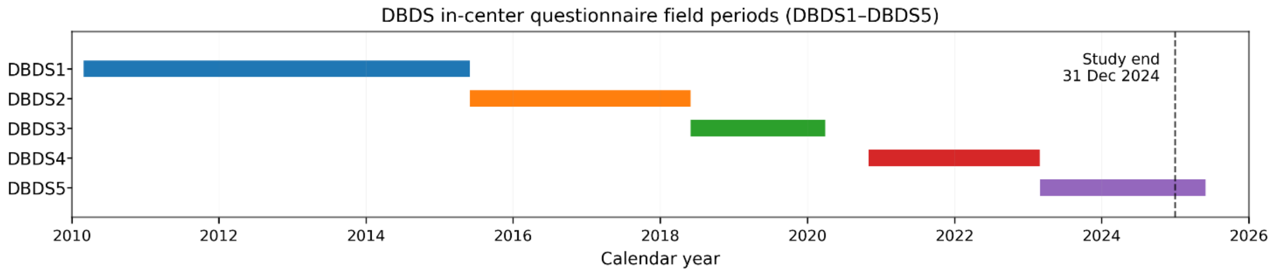
